# Afterload-Adjusted Strain Refines Risk Stratification in Aortic Stenosis With Varying Degrees of Aortic Regurgitation

**DOI:** 10.1101/2025.10.25.25338684

**Authors:** Jonghee Sun, Jiesuck Park, MinJung Bak, Hong-Mi Choi, In-Chang Hwang, Yeonyee E. Yoon, Goo-Yeong Cho

**Author notes:** **Address for correspondence:** Jiesuck Park, MD, Department of Cardiology, Cardiovascular Center Seoul National University Bundang Hospital 82, Gumi-ro 173beon-gil, Seongnam, Gyeonggi-do, 13620 Republic of Korea.

## Abstract

**Background and Objectives:** In aortic stenosis (AS), concomitant aortic regurgitation (AR) may cause heterogeneous left ventricular (LV) remodeling and myocardial responses beyond valve-centric assessment. We evaluated LV geometry and myocardial deformation relative to systolic pressure load in patients with AS across varying AR severity.

**Methods:** We analyzed 950 patients with moderate or greater AS categorized as no AR, mild AR, or moderate or greater AR. LV geometry was assessed using LV mass index (LVMI) and relative wall thickness (RWT). Afterload-adjusted strain (AAS) was calculated as absolute LV global longitudinal strain (LVGLS) multiplied by estimated LV systolic pressure. The primary outcome was all-cause death or heart failure hospitalization.

**Results:** During a median follow-up of 3.5 years, 122 primary events occurred. Increasing AR severity was associated with LV dilation, increased LVMI, and reduced RWT, whereas AAS remained similar across AR categories. LV geometric measures were not independently associated with the primary outcome. Lower AAS was independently associated with the primary outcome (adjusted HR 1.33 per 500-unit decrease, 95% CI 1.12–1.58; p=0.001; adjusted HR 1.41 per 1-SD decrease, 95% CI 1.14–1.73; p=0.001). AAS provided incremental prognostic value beyond clinical and LV geometric variables, increasing the 1-year AUC from 0.762 to 0.788 (p<0.001).

**Conclusions:** Among patients with moderate or greater AS across varying degrees of concomitant AR, increasing AR severity was associated with progressive LV remodeling, whereas lower AAS was independently associated with adverse outcomes. Relating myocardial deformation to systolic pressure load may refine risk stratification beyond clinical and LV geometric measures.

**Author’s Summary:** In patients with moderate or greater aortic stenosis and concomitant aortic regurgitation, valve-centric assessment may not fully characterize the heterogeneous ventricular response to combined pressure and volume loading. We assessed left ventricular geometry using LV mass index and relative wall thickness and evaluated afterload-adjusted strain (AAS), which integrates myocardial deformation with the prevailing systolic pressure load. Increasing aortic regurgitation severity was associated with progressive geometric remodeling, whereas lower AAS was independently associated with death or heart failure hospitalization. AAS also improved prognostic discrimination beyond clinical and geometric variables, supporting a more comprehensive, myocardium-centered approach to risk assessment.

## 1. INTRODUCTION

In patients with aortic stenosis (AS), chronic pressure overload promotes concentric hypertrophic remodeling, whereas concomitant aortic regurgitation (AR) introduces an additional volume-loading component that may lead to left ventricular (LV) dilation and alter the resultant geometric phenotype.^1–3^ The interaction between these loading conditions may produce heterogeneous patterns of LV remodeling and myocardial response across increasing degrees of concomitant AR.^1–3^ Despite this complex pathophysiology, echocardiographic assessment remains largely valve-centric, focusing primarily on the severity of AS and AR.^4^ Conventional valve parameters may therefore incompletely characterize the ventricular consequences of combined pressure and volume loading.^5^ Recent studies have shown that LV remodeling and myocardial dysfunction are important determinants of clinical outcomes in patients with AS or AR,^6,7^ highlighting the need to assess the ventricular response beyond valve severity alone.^8^

Accordingly, we evaluated an integrated echocardiographic framework incorporating two complementary dimensions of ventricular response: LV geometric remodeling assessed using conventional structural measures, including LV mass index (LVMI) and relative wall thickness (RWT), and myocardial deformation evaluated in relation to the total systolic pressure load. Afterload-adjusted strain (AAS) was constructed as an integrated pressure– deformation index combining LV global longitudinal strain (LVGLS) with estimated LV systolic pressure derived from systolic blood pressure (SBP) and aortic valve mean pressure gradient (AV mPG). Using this framework, we aimed to characterize changes in LV geometry across increasing degrees of concomitant AR and to determine whether AAS provides prognostic information beyond conventional clinical, geometric, and strain-based measures.

## 2. METHODS

### 2.1. Study Population

We retrospectively screened 4,477 consecutive patients who underwent transthoracic echocardiography (TTE) at Seoul National University Bundang Hospital between January 1, 2015, and December 31, 2020, and were identified as having AS or AR. Patients with mild AS were excluded to focus on clinically relevant valvular loading conditions. Additional exclusion criteria included concomitant moderate or greater mitral stenosis or regurgitation, prior mitral or aortic valve surgery, bicuspid aortic valve, known cardiomyopathy, LV ejection fraction (LVEF) <40%, atrial fibrillation or flutter, cardiac implantable electronic devices, left bundle branch block, uncontrolled hypertension (systolic blood pressure >180 mmHg), severe pulmonary hypertension, poor echocardiographic image quality, or incomplete clinical or echocardiographic data. After applying these criteria, 950 patients with moderate or greater AS were included in the final cohort. Patients were further categorized according to the presence and severity of concomitant AR into three groups: (1) moderate or greater AS without AR, (2) moderate or greater AS with mild AR, and (3) moderate or greater AS with moderate or greater AR. The study flow and patient selection process are summarized in Figure 1. The study protocol complied with the Declaration of Helsinki and was approved by the Institutional Review Board of Seoul National University Bundang Hospital (IRB No. B-2511-1008-105). The requirement for informed consent was waived due to the retrospective nature of the study.

**Figure 1.**
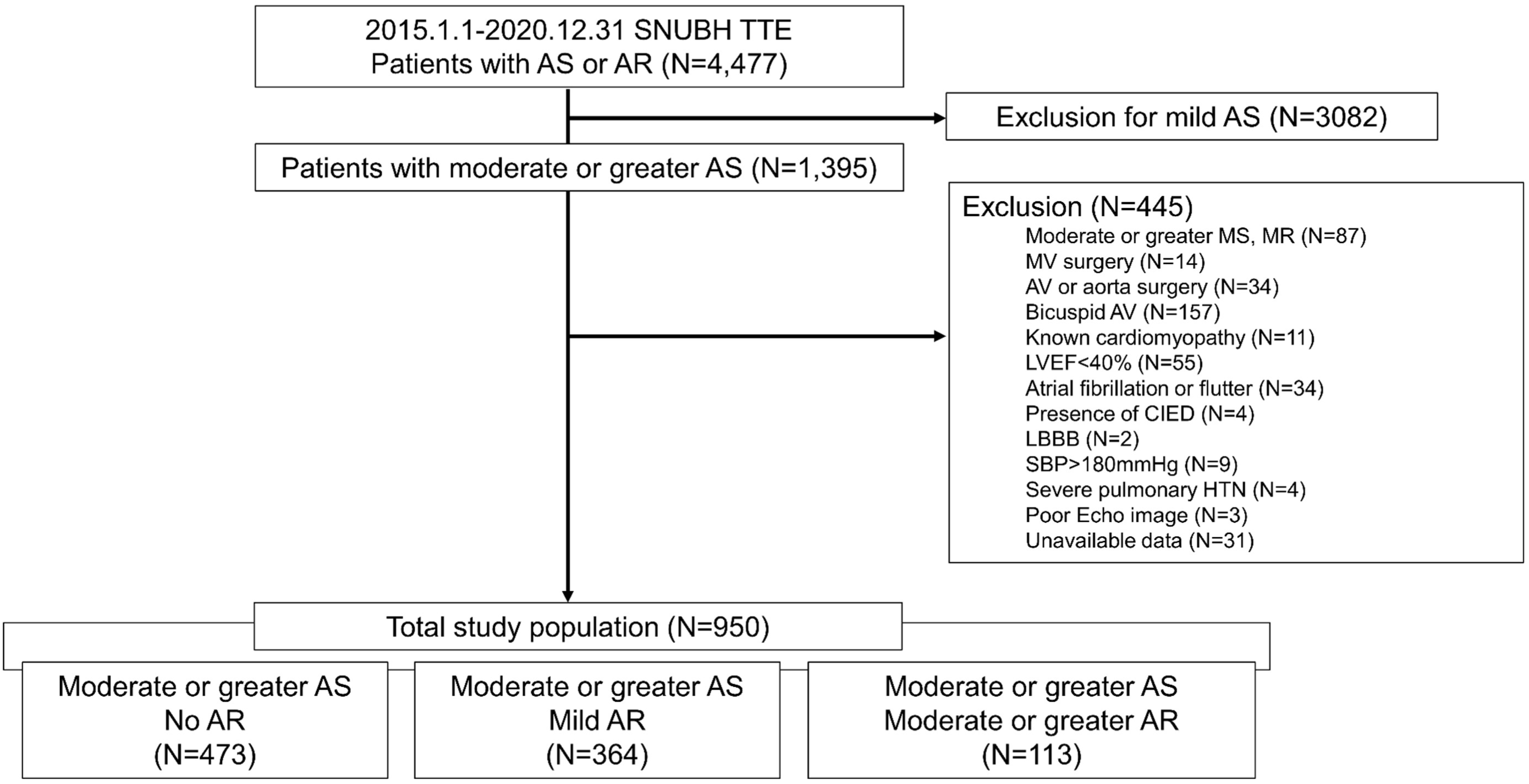
Study flow diagram. Patients were screened from consecutive transthoracic echocardiography studies at Seoul National University Bundang Hospital between January 2015 and December 2020. After applying exclusion criteria, 950 patients with moderate or greater AS were categorized according to concomitant AR severity. CIED = cardiac implantable electronic device; LBBB = left bundle branch block; LVEF = left ventricular ejection fraction; MR = mitral regurgitation; MS = mitral stenosis; SBP = systolic blood pressure.

### 2.2. Echocardiographic Assessment

All echocardiographic examinations were performed by trained echocardiographers and interpreted by board-certified cardiologists according to contemporary guideline recommendations as part of routine clinical care.^9^ Blood pressure was measured at the time of the echocardiographic examination as part of the routine institutional protocol, using an automated oscillometric sphygmomanometer applied to the upper arm. The contemporaneously measured SBP was used for the calculation of AAS. The severity of AS and AR was determined from the final echocardiographic interpretation based on the available Doppler, quantitative, and qualitative findings. AS severity was assessed using peak aortic jet velocity (AV Vmax), mean transaortic pressure gradient (AV mPG), and aortic valve area (AVA) calculated by the continuity equation.^4^ AR severity was determined using an integrative approach based on available parameters, including color Doppler flow convergence, jet density, deceleration rate, diastolic flow reversal in the descending aorta, and vena contracta width.^10^ Cardiac chamber quantification included LV end-diastolic diameter, interventricular septal thickness, posterior wall thickness, RWT, LV mass index (LVMI), LVEDVi, and LVEF. LV volumes and LVEF were calculated using the biplane Simpson method. Diastolic function was assessed using transmitral Doppler inflow and tissue Doppler imaging, including the E/e′ ratio. LVGLS was measured using two-dimensional speckle-tracking echocardiography with dedicated software (TomTec Image Arena 4.6, TomTec Imaging Systems, Munich, Germany; or EchoPAC PC BT20, GE Medical Systems, Horten, Norway). For the calculation of AAS, LVGLS was entered as an absolute value.^11,12^

### 2.3. Integrated Hemodynamic Assessment

To characterize the ventricular response to the combined hemodynamic burden of aortic valve disease, we used an integrated echocardiographic framework incorporating two complementary components: LV geometric remodeling assessed using conventional structural parameters, and myocardial performance evaluated in relation to the total systolic pressure load (**Figure 2**).

**Figure 2.**
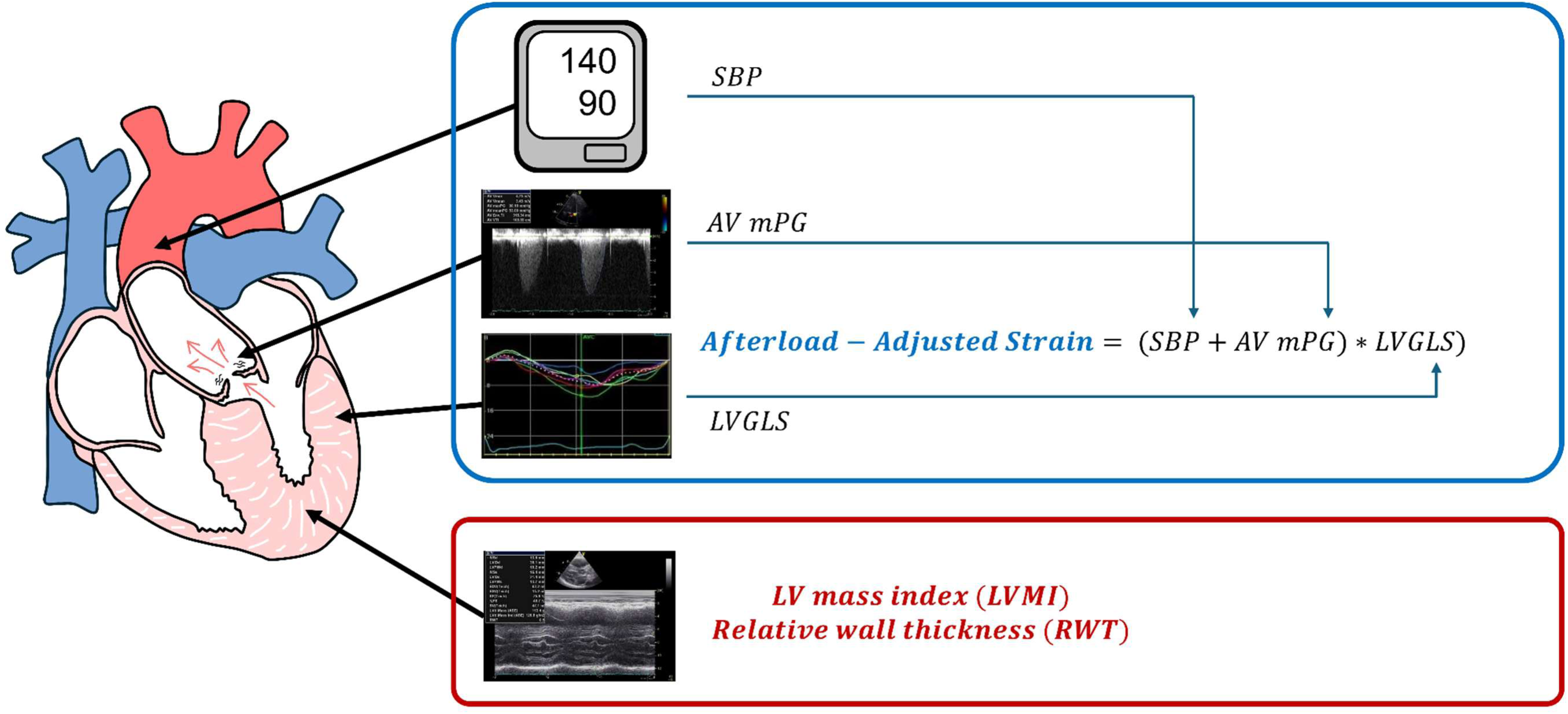
Integrated echocardiographic framework. The integrated echocardiographic framework incorporates two complementary dimensions of ventricular response: left ventricular geometric remodeling assessed using LV mass index (LVMI) and relative wall thickness (RWT), and myocardial deformation evaluated in relation to systolic pressure load using afterload-adjusted strain (AAS). AAS is calculated as |LVGLS| × (SBP + AV mPG). Higher AAS values indicate greater myocardial deformation under the prevailing systolic pressure load. AAS = afterload-adjusted strain; AV mPG = aortic valve mean pressure gradient; LV = left ventricle; LVGLS = left ventricular global longitudinal strain; LVMI = left ventricular mass index; RWT = relative wall thickness; SBP = systolic blood pressure.

#### 2.3.1. Left Ventricular Geometric Remodeling

LV geometric remodeling was characterized using LV mass index (LVMI) and relative wall thickness (RWT). LVMI reflects the extent of LV hypertrophic remodeling, whereas RWT characterizes the LV geometric pattern, particularly concentric remodeling. Both parameters were evaluated separately as established and clinically interpretable measures of LV geometry.

#### 2.3.2. Afterload-Adjusted Strain

LV myocardial deformation was assessed using LVGLS obtained from two-dimensional speckle-tracking echocardiography. Because LVGLS is load-dependent,^12^ AAS was constructed as an integrated pressure–deformation index that relates myocardial deformation to the total systolic pressure load imposed on the LV, rather than directly normalizing strain for afterload. Effective LV systolic pressure was approximated as the sum of systolic blood pressure (SBP) and AV mPG based on a previously described approach in patients with AS.^13^ AAS was calculated as:

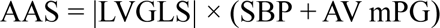

where LVGLS is expressed as an absolute value (%) and SBP and AV mPG are expressed in mmHg. Higher AAS values indicate greater myocardial deformation under the prevailing systolic pressure load, whereas lower values indicate impaired myocardial performance in relation to that load.

### 2.4. Ascertainment of Clinical Information and Outcome Definitions

Baseline clinical and laboratory data were ascertained through systematic review of the institutional electronic medical record. For hemoglobin, albumin, serum creatinine, and estimated glomerular filtration rate (eGFR), the measurements closest to the index TTE were used. The primary outcome was a composite of all-cause death or hospitalization for heart failure (HHF). The secondary outcome was aortic valve replacement (AVR), including both surgical and transcatheter interventions. Clinical events were identified through review of institutional hospital records. Follow-up duration was defined from the index TTE to the first occurrence of an outcome event or the last available clinical contact, whichever occurred first.

### 2.5. Statistical Analysis

Clinical and echocardiographic characteristics were compared according to concomitant AR categories. Continuous variables were presented as median (interquartile range), and categorical variables as counts (percentages). Group comparisons were performed using the Kruskal–Wallis test for continuous variables and the χ² test for categorical variables. The distributions of LVEDVi, RWT, LVMI, LVEF, SBP, AV mPG, LVGLS, and AAS across concomitant AR categories were illustrated using box-and-whisker plots. For variables showing a significant overall Kruskal–Wallis test, post hoc pairwise comparisons were performed using Dunn’s test with Bonferroni correction.

AAS was analyzed as a continuous variable, with hazard ratios reported per 500-unit decrease in the raw value and per 1-SD decrease. LVMI and RWT were evaluated separately as measures of LV geometric remodeling. For categorical and subgroup analyses, AAS was dichotomized using an optimal cutoff of 2,390, determined by the Youden index. LV hypertrophy (LVH) was defined as an LVMI >115 g/m² in men or >95 g/m² in women, and increased RWT was defined as RWT >0.42. Based on LVH status and RWT, conventional LV remodeling patterns were classified according to established geometric criteria.^9^ Reduced LVGLS was defined as an absolute value <15.9%, corresponding to the cohort median.

Cumulative event probability for the primary outcome was estimated using the Kaplan–Meier method and compared using the log-rank test. Associations between echocardiographic indices and clinical outcomes were evaluated using Cox proportional hazards regression models. Multivariable models were adjusted for age, sex, LVEF, LVMI, RWT, LAVI, E/e′, hypertension, diabetes mellitus, eGFR, hemoglobin, and albumin. A joint model including both AAS and LVGLS was additionally constructed to assess whether AAS provided prognostic information beyond LVGLS. Restricted cubic spline analyses were performed to examine the continuous associations of AAS, LVMI, and RWT with the primary and secondary outcomes and to assess potential non-linearity. The spline models used the same covariate adjustment as the corresponding multivariable Cox regression models.

Additional outcome analyses were performed for the individual components of the primary outcome (all-cause death and HHF) and for the cardiovascular-specific composite outcome of cardiovascular death or HHF.

To evaluate whether the prognostic association of AAS was preserved among patients with concomitant AR, a sensitivity analysis was performed after restricting the cohort to patients with at least mild AR. Because AVR may modify the subsequent risk of death or HHF, several sensitivity analyses were performed to assess the influence of AVR during follow-up. These included a Fine–Gray analysis treating AVR as a competing event, an extended Cox model incorporating AVR as a time-dependent covariate, and an analysis censoring follow-up at the time of AVR. A post-AVR landmark analysis was additionally performed among patients who underwent AVR and had follow-up beyond the intervention date.

The incremental prognostic value of AAS was assessed by constructing receiver operating characteristic (ROC) curves for the 1-year risk of the primary outcome using sequential models. The 1-year horizon was selected to represent a clinically relevant near-term period for risk stratification. The baseline model included age, sex, LVEF, LAVI, E/e’, hypertension, diabetes mellitus, eGFR, hemoglobin, and albumin. LVMI and RWT were subsequently added to the baseline model, followed by AAS. Discriminative performance was evaluated using the area under the curve (AUC) and Harrell’s C-index for the 1-year risk of the primary outcome. Differences in AUC between nested models were compared using the DeLong test. Incremental prognostic performance was further assessed using continuous net reclassification improvement (NRI) and integrated discrimination improvement (IDI). Clinical utility was evaluated using decision-curve analysis across threshold probabilities ranging from 2% to 15% for the predicted 1-year risk of the primary outcome. Cox model–predicted 1-year risks according to AAS were also estimated by sex, with continuous covariates fixed at their cohort median values and binary covariates fixed at their most frequent categories. All statistical analyses were performed using R software (version 4.5.1; R Foundation for Statistical Computing, Vienna, Austria). A two-sided p value <0.05 was considered statistically significant.

## 3. RESULTS

### 3.1. Baseline Characteristics According to Concomitant AR Severity

Baseline clinical and echocardiographic characteristics according to concomitant AR severity are summarized in **Table 1**. Clinical characteristics were largely comparable across groups, although patients with moderate or greater AR tended to be younger and had a lower prevalence of diabetes. Both AV Vmax and AV mPG increased with increasing AR severity (both p<0.001), whereas AVA did not differ significantly among groups. With increasing AR severity, LV geometry progressively shifted from a concentric remodeling pattern toward a more dilated pattern, characterized by larger LVEDD and LVEDVi, increased LVMI, and decreased RWT, despite no significant change in LVEF. LVMI increased and RWT decreased progressively with increasing AR severity (both p<0.001), reflecting distinct changes in LV geometric remodeling. In contrast, SBP and LVGLS did not differ significantly across concomitant AR categories. Accordingly, AAS remained comparable across groups (p=0.649; **Figure 3**).

**Figure 3.**
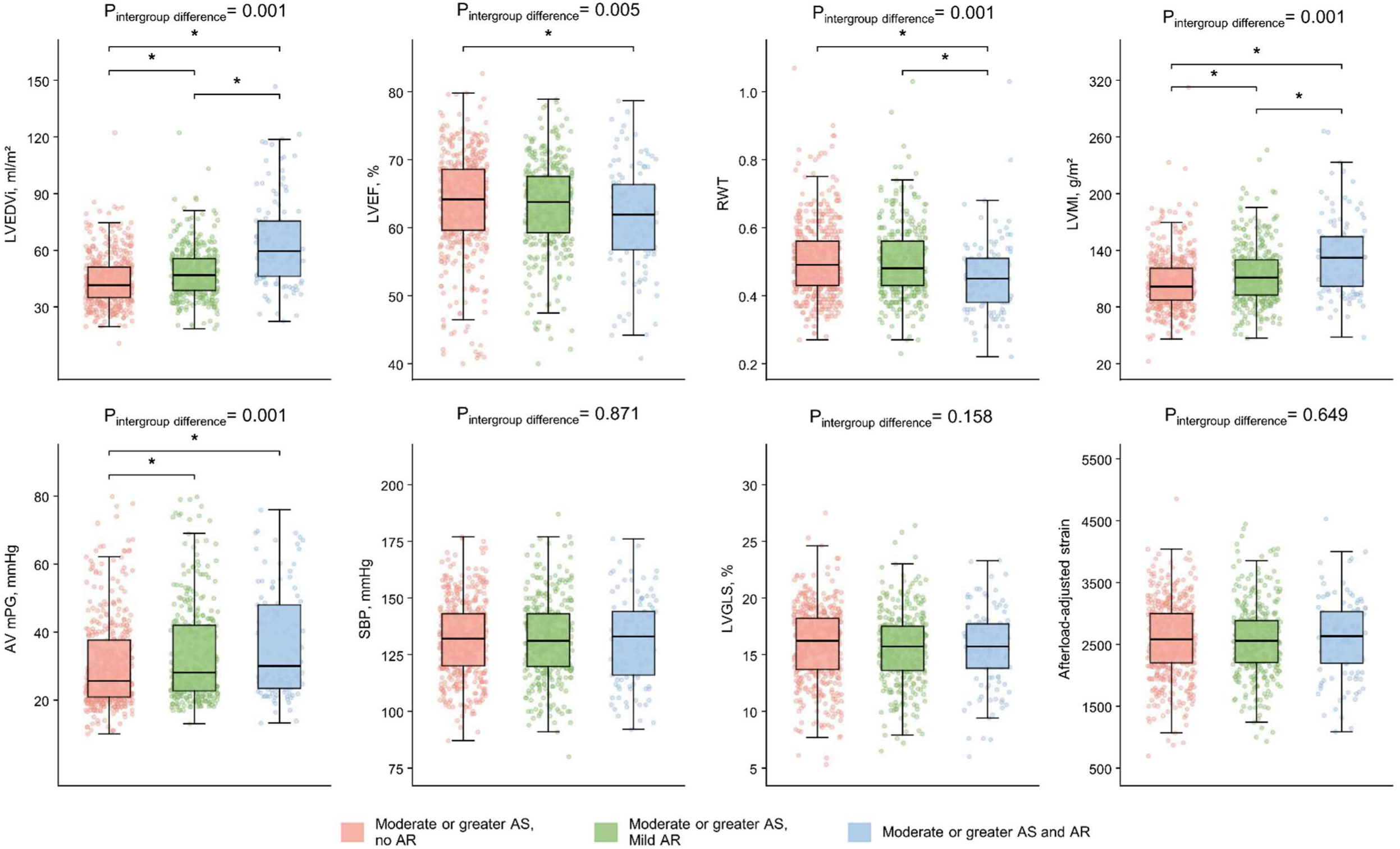
Distribution of echocardiographic parameters according to concomitant aortic regurgitation severity. Box-and-whisker plots show LVEDVi, RWT, LVMI, LVEF, SBP, AV mPG, LVGLS, and AAS across concomitant AR categories. Overall group differences were tested using the Kruskal–Wallis test. For variables with significant overall differences, pairwise post hoc comparisons were performed using Dunn’s test with Bonferroni correction (*p < 0.05). AAS = afterload-adjusted strain; AR = aortic regurgitation; AV mPG = aortic valve mean pressure gradient; LVEDVi = indexed left ventricular end-diastolic volume; LVEF = left ventricular ejection fraction; LVGLS = left ventricular global longitudinal strain; LVMI = left ventricular mass index; RWT = relative wall thickness; SBP = systolic blood pressure.

**Table 1.** Baseline characteristics according to concomitant aortic regurgitation severity.

| **Variables** | **Overall population**<br>**(N=950)** | **Moderate≤ AS,**<br>**No AR**<br>**(N=473)** | **Moderate≤ AS,**<br>**Mild AR**<br>**(N=364)** | **Moderate≤ AS,**<br>**Moderate≤ AR**<br>**(N=113)** | **p** |
| --- | --- | --- | --- | --- | --- |
| Clinical variables |  |  |  |  |  |
| Age, years | 80 (74–85) | 81 (75–85) | 80 (73–84) | 78 (70–84) | 0.003 |
| Male | 434 (45.7) | 213 (45.0) | 174 (47.8) | 47 (41.6) | 0.472 |
| BMI, kg/m<sup>2</sup> | 24.2 (21.8–26.7) | 24.2 (22–26.8) | 24.3 (21.8–26.7) | 23.6 (21.1–26.1) | 0.155 |
| SBP, mmHg | 132 (120–143) | 132 (120–143) | 131 (120–143) | 133 (116–144) | 0.871 |
| DBP, mmHg | 71 (64–78) | 72 (65–80) | 70 (63–78) | 67 (58–77) | 0.001 |
| Hypertension | 202 (21.3) | 105 (22.2) | 80 (22.0) | 17 (15.0) | 0.227 |
| Diabetes | 177 (18.6) | 99 (20.9) | 66 (18.1) | 12 (10.6) | 0.039 |
| Myocardial infarction | 43 (4.5) | 24 (5.1) | 14 (3.8) | 5 (4.4) | 0.697 |
| Congestive heart failure | 93 (9.8) | 42 (8.9) | 37 (10.2) | 14 (12.4) | 0.505 |
| Stroke | 101 (10.6) | 58 (12.3) | 34 (9.3) | 9 (8.0) | 0.246 |
| Chronic kidney disease | 84 (8.8) | 39 (8.2) | 49 (10.7) | 6 (5.3) | 0.170 |
| Laboratory |  |  |  |  |  |
| Hemoglobin, g/dL | 12.0 (10.7–13.2) | 12.0 (10.6–13.3) | 12.0 (10.7–13.1) | 12.0 (10.8–13.1) | 0.713 |
| Serum albumin, g/dL | 3.9 (3.5–4.2) | 3.9 (3.4–4.2) | 3.9 (3.6–4.2) | 3.9 (3.6–4.1) | 0.644 |
| Serum creatinine, mg/dL | 0.90 (0.71–1.19) | 0.88 (0.71–1.14) | 0.95 (0.73–1.27) | 0.87 (0.70–1.17) | 0.055 |
| eGFR (CKD-EPI), mL/min/1.73m<sup>2</sup> | 71.3 (50.6–84.6) | 71.9 (53.2–84.6) | 67.8 (45.4–84.2) | 72.8 (52.7–86.4) | 0.081 |
| MAVD severity |  |  |  |  |  |
| AS grade |  |  |  |  | 0.058 |
| Moderate | 604(63.6) | 344 (72.7) | 234 (64.3) | 74 (65.5) |  |
| Severe | 346 (36.4) | 129 (27.3) | 130 (35.7) | 39 (34.5) |  |
| AR grade |  |  |  |  | 0.001 |
| No AR | 473 (49.8) | 473 (100) | 0 (0.0) | 0 (0.0) |  |
| Mild | 364 (38.3) | 0 (0.0) | 364 (100.0) | 0 (0.0) |  |
| Moderate | 97 (10.2) | 0 (0.0) | 0 (0.0) | 97 (85.8) |  |
| Severe | 16 (1.7) | 0 (0.0) | 0 (0.0) | 16 (14.2) |  |
| Echocardiography |  |  |  |  |  |
| IVS, mm | 12 (10–13) | 12 (10–13) | 12 (11–13) | 12 (10–13) | 0.930 |
| PW, mm | 11 (10–12) | 10 (9–12) | 11 (10–12) | 11 (9–12) | 0.166 |
| LVEDD, mm | 44 (40–49) | 43 (39–47) | 45 (41–49) | 49 (43–54) | 0.001 |
| RWT | 0.5 (0.4–0.6) | 0.5 (0.4–0.6) | 0.5 (0.4–0.6) | 0.4 (0.4–0.5) | 0.001 |
| LVEDV, ml | 73 (59–92) | 67 (54–86) | 76 (61–92) | 92 (74–129) | 0.001 |
| LVEDVi, ml/m<sup>2</sup> | 45 (37–56) | 42 (35–51) | 47 (39–56) | 60 (46–76) | 0.001 |
| LVEF, % | 64 (59–68) | 64 (60–69) | 64 (59–68) | 62 (57–66) | 0.005 |
| LAVI, ml/m<sup>2</sup> | 48 (38–60) | 46 (37–57) | 48 (38–61) | 55 (43–69) | 0.001 |
| LVMI, g/m<sup>2</sup> | 107 (91–129) | 102 (87–121) | 111 (93–130) | 132 (102–155) | 0.001 |
| AV Vmax, m/s | 3.5 (3.2–4.2) | 3.4 (3.1– 4.1) | 3.6 (3.2– 4.3) | 3.8 (3.4–4.6) | 0.001 |
| AV mPG, mmHg | 27 (22–41) | 26 (21–38) | 28 (23–42) | 30 (23–48) | 0.001 |
| AVA, cm<sup>2</sup> | 1.1 (0.9–1.3) | 1.2 (0.9–1.3) | 1.1 (0.9– 1.3) | 1.1 (0.8–1.3) | 0.161 |
| E/e' | 16 (12–21) | 16 (12–21) | 16 (12–21) | 17 (13–23) | 0.485 |
| TR Vmax, m/s | 2.6 (2.3–2.9) | 2.6 (2.3– 2.8) | 2.6 (2.3–2.9) | 2.6 (2.4–2.9) | 0.553 |
| RVSP, mmHg | 32 (26–39) | 32 (26–39) | 32 (26–39) | 32 (28–41) | 0.381 |
| LVGLS, % | 16 (14–18) | 16 (14–18) | 16 (14–18) | 16 (14–18) | 0.158 |
| AAS | 2572 (2204–2961) | 2579 (2200–2997) | 2553 (2205–2886) | 2629 (2194–3028) | 0.649 |

Values are presented as median (interquartile range) or number (percentage), unless otherwise indicated.
AAS, afterload-adjusted strain; AR, aortic regurgitation; AS, aortic stenosis; AVA, aortic valve area; AV mPG, mean transaortic pressure gradient; AV Vmax, peak aortic jet velocity; BMI, body mass index; DBP, diastolic blood pressure; E/e', ratio of early transmitral flow velocity to early diastolic mitral annular velocity; IVS, interventricular septal thickness; LAVI, left atrial volume index; LVEDD, left ventricular end-diastolic diameter; LVEDV, left ventricular end-diastolic volume; LVEDVi, indexed left ventricular end-diastolic volume; LVEF, left ventricular ejection fraction; LVGLS, left ventricular global longitudinal strain; LVMI, left ventricular mass index; PW, posterior wall thickness; RVSP, right ventricular systolic pressure; RWT, relative wall thickness; SBP, systolic blood pressure; TR Vmax, peak tricuspid regurgitation velocity.

### 3.2. Clinical Outcomes According to Concomitant AR Severity and AAS

During a median follow-up of 3.5 years (IQR 0.6–5.6), a total of 122 primary outcome events occurred, including 75 all-cause deaths and 47 HHF events. The cumulative event probability of the primary outcome did not differ significantly across concomitant AR categories, LVH status, or RWT category. In contrast, patients with low AAS had a significantly higher cumulative incidence of the primary outcome than those with high AAS (log-rank p<0.001) (**Figure 4A**).

**Figure 4.**
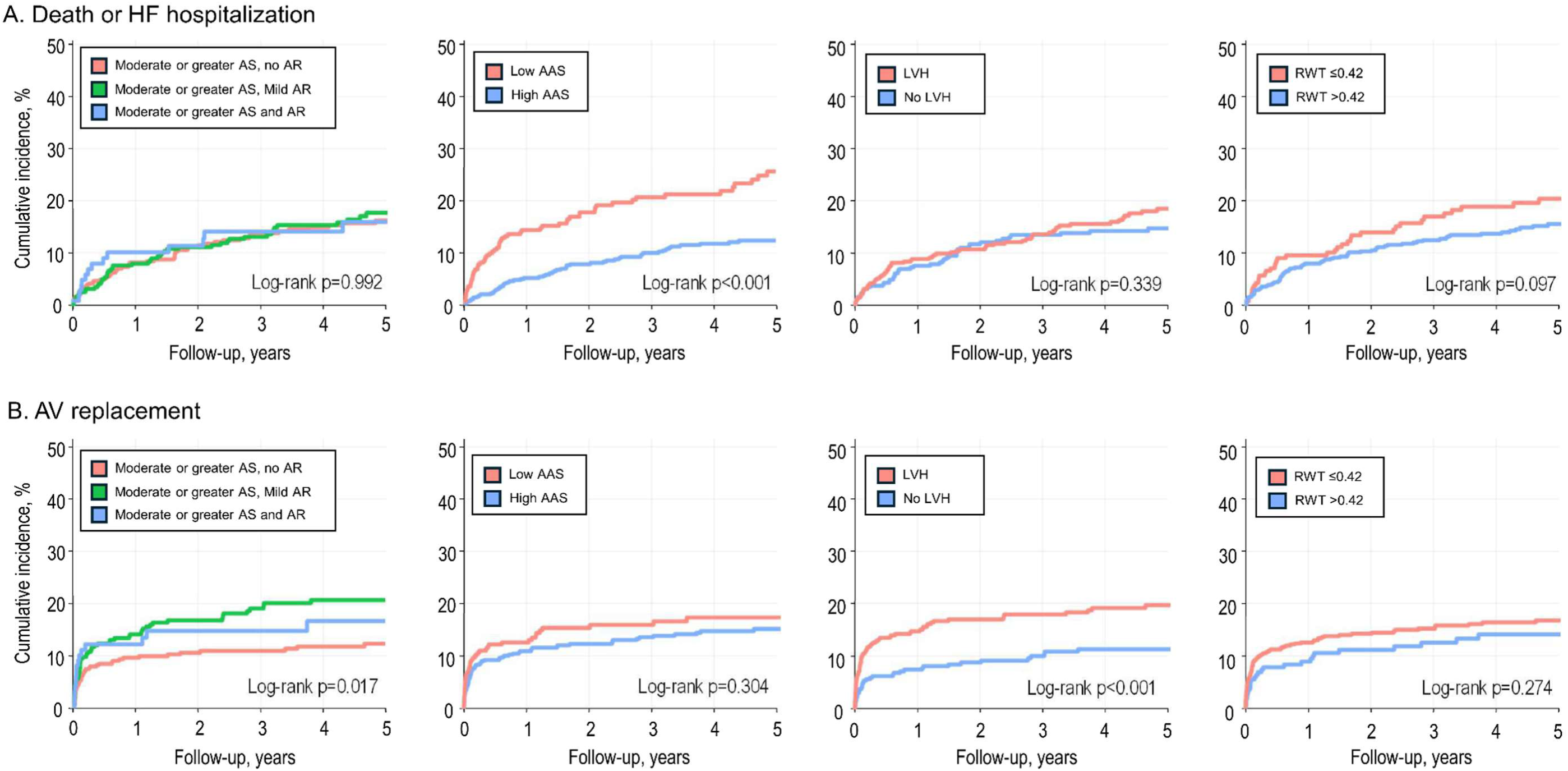
Kaplan–Meier curves for cumulative event probabilities. Kaplan–Meier curves showing cumulative event probabilities for (A) the primary outcome of all-cause death or heart failure hospitalization and (B) the secondary outcome of aortic valve replacement, stratified according to concomitant AR category, LV hypertrophy status, RWT category, and AAS status. AAS was dichotomized using a cutoff of 2,390. AAS = afterload-adjusted strain; AR = aortic regurgitation; AVR = aortic valve replacement; LVH = left ventricular hypertrophy; RWT = relative wall thickness.

In Cox proportional hazards analyses, concomitant AR categories and conventional LV geometric parameters were not independently associated with the primary outcome (**Table 2**). By contrast, lower AAS remained independently associated with a higher risk of the primary outcome, with an adjusted HR of 1.33 per 500-unit decrease (95% CI 1.12–1.58; p=0.001) and 1.41 per 1-SD decrease (95% CI 1.14–1.73; p=0.001). Similarly, LVGLS was independently associated with the primary outcome when evaluated in a separate model (adjusted HR 1.08 per 1% decrease, 95% CI 1.01–1.15; p=0.022). However, when both indices were included in a joint model, AAS remained independently associated with the primary outcome (adjusted HR 1.44 per 500-unit decrease, 95% CI 1.07–1.94; p=0.016), whereas LVGLS was no longer significant (adjusted HR 0.97 per 1% decrease, 95% CI 0.87– 1.07; p=0.522). Consistent with these findings, restricted cubic spline analysis demonstrated a graded increase in the risk of the primary outcome with decreasing AAS, whereas LVMI and RWT showed no significant associations with the primary outcome (**Figure S1A**). Subgroup analyses showed that the association between lower AAS and the primary outcome was generally consistent across concomitant AR categories, LVH status, RWT category, and the presence or absence of prior myocardial infarction or heart failure, with no significant interactions (**Figure 5**). The association was directionally stronger in patients with reduced LVGLS, although the interaction according to LVGLS status was not statistically significant.

**Figure 5.**
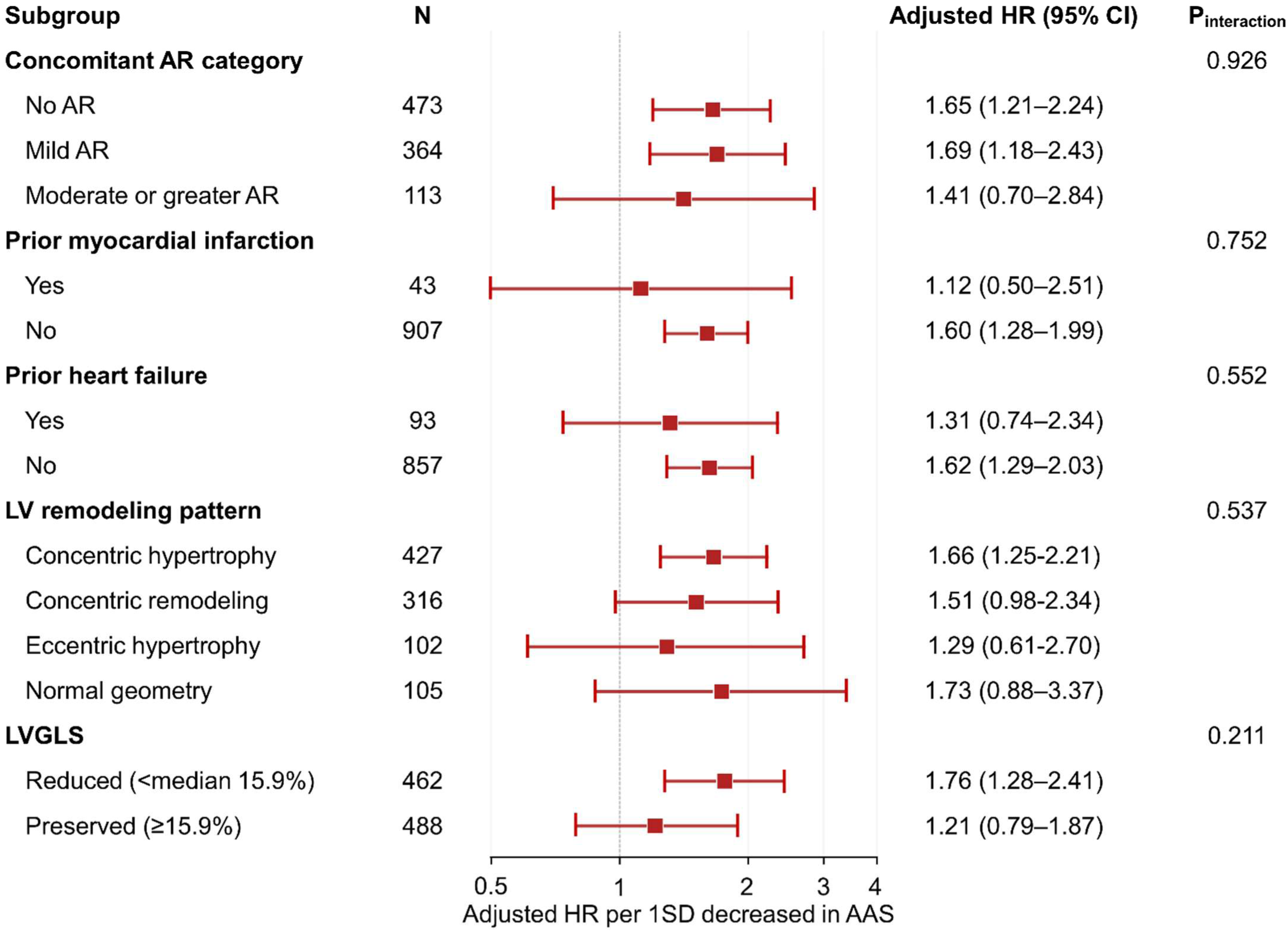
Subgroup analysis for the primary outcome. Forest plot showing adjusted hazard ratios (HRs) per 1-SD decrease in AAS for the primary outcome across concomitant AR categories and subgroups defined by prior myocardial infarction, prior heart failure, conventional LV remodeling pattern, and LVGLS strata. Conventional LV remodeling patterns were classified as concentric hypertrophy, concentric remodeling, eccentric hypertrophy, or normal geometry according to LVMI and RWT. Reduced LVGLS was defined as an absolute LVGLS value <15.9%, corresponding to the cohort median. AAS = afterload-adjusted strain; AR = aortic regurgitation; HR = hazard ratio; LV = left ventricle; LVGLS = left ventricular global longitudinal strain; LVMI = left ventricular mass index; RWT = relative wall thickness; SD = standard deviation.

**Table 2.**
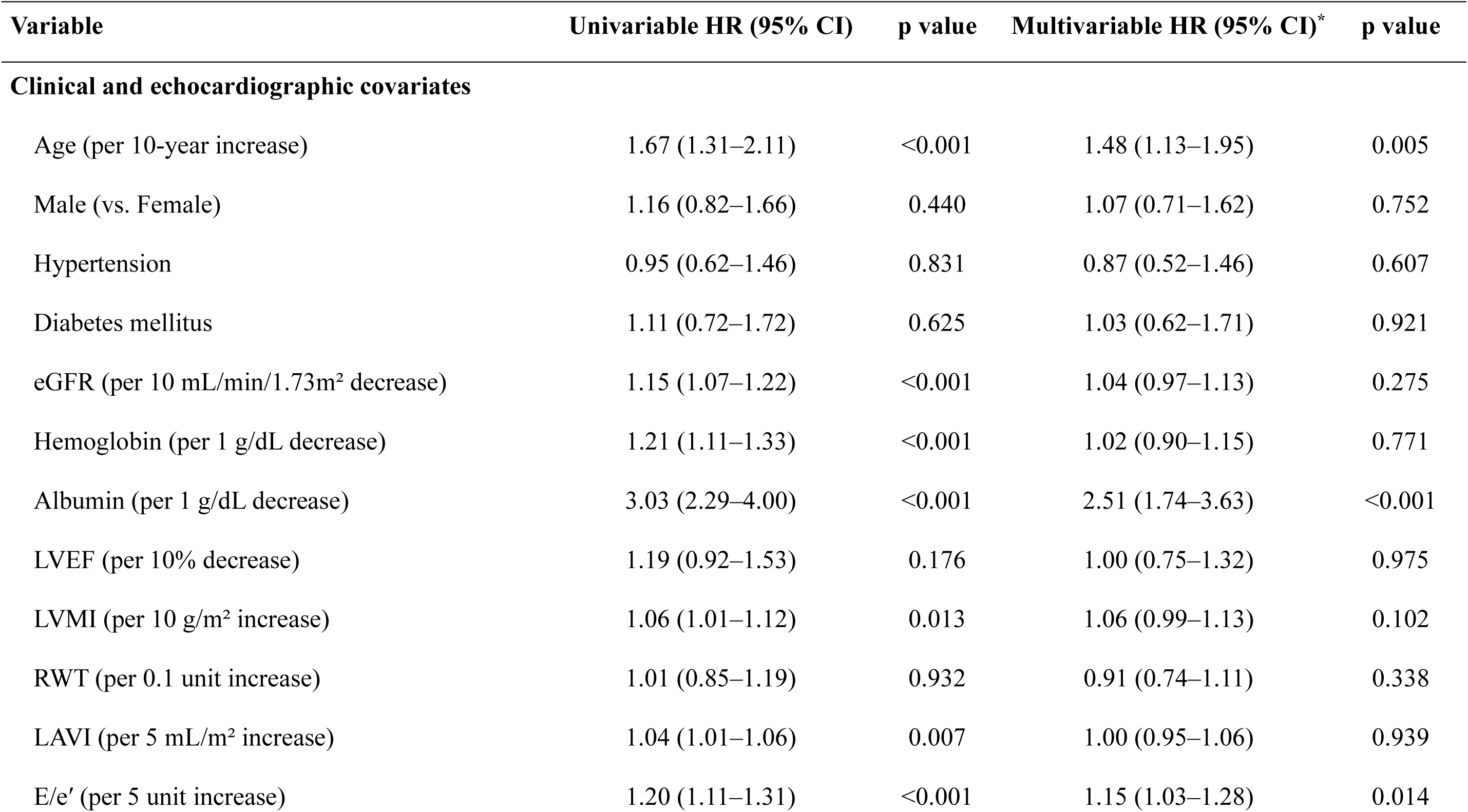
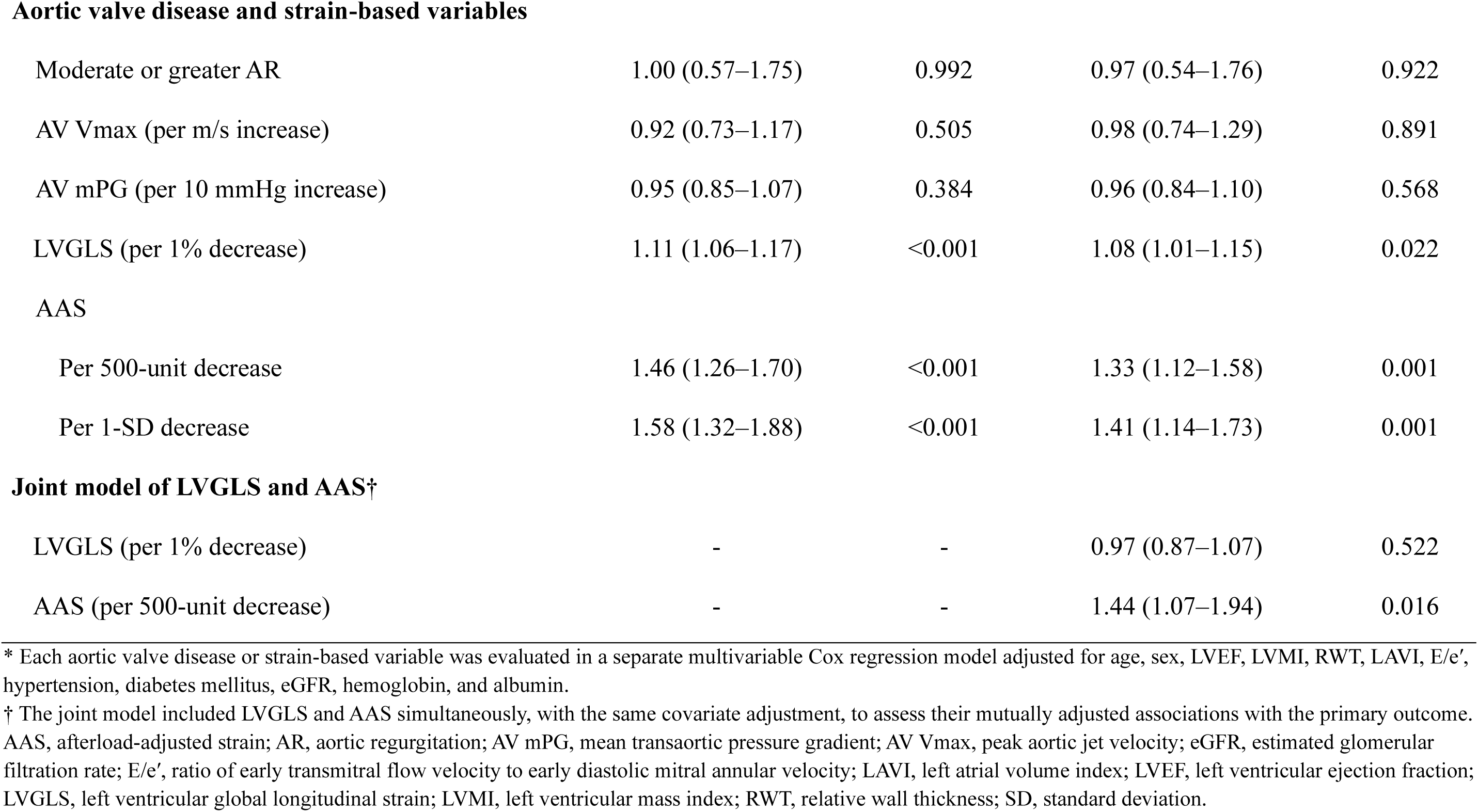
Cox regression analyses for the primary outcome.

| Variable | Univariable HR (95% CI) | p value | Multivariable HR (95% CI)* | p value |
| --- | --- | --- | --- | --- |
| **Clinical and echocardiographic covariates** |  |  |  |  |
| Age (per 10-year increase) | 1.67 (1.31–2.11) | <0.001 | 1.48 (1.13–1.95) | 0.005 |
| Male (vs. Female) | 1.16 (0.82–1.66) | 0.440 | 1.07 (0.71–1.62) | 0.752 |
| Hypertension | 0.95 (0.62–1.46) | 0.831 | 0.87 (0.52–1.46) | 0.607 |
| Diabetes mellitus | 1.11 (0.72–1.72) | 0.625 | 1.03 (0.62–1.71) | 0.921 |
| eGFR (per 10 mL/min/1.73m<sup>2</sup> decrease) | 1.15 (1.07–1.22) | <0.001 | 1.04 (0.97–1.13) | 0.275 |
| Hemoglobin (per 1 g/dL decrease) | 1.21 (1.11–1.33) | <0.001 | 1.02 (0.90–1.15) | 0.771 |
| Albumin (per 1 g/dL decrease) | 3.03 (2.29–4.00) | <0.001 | 2.51 (1.74–3.63) | <0.001 |
| LVEF (per 10% decrease) | 1.19 (0.92–1.53) | 0.176 | 1.00 (0.75–1.32) | 0.975 |
| LVMI (per 10 g/m<sup>2</sup> increase) | 1.06 (1.01–1.12) | 0.013 | 1.06 (0.99–1.13) | 0.102 |
| RWT (per 0.1 unit increase) | 1.01 (0.85–1.19) | 0.932 | 0.91 (0.74–1.11) | 0.338 |
| LAVI (per 5 mL/m<sup>2</sup> increase) | 1.04 (1.01–1.06) | 0.007 | 1.00 (0.95–1.06) | 0.939 |
| E/e' (per 5 unit increase) | 1.20 (1.11–1.31) | <0.001 | 1.15 (1.03–1.28) | 0.014 |
| Aortic valve disease and strain-based variables |  |  |  |  |
| Moderate or greater AR | 1.00 (0.57–1.75) | 0.992 | 0.97 (0.54–1.76) | 0.922 |
| AV Vmax (per m/s increase) | 0.92 (0.73–1.17) | 0.505 | 0.98 (0.74–1.29) | 0.891 |
| AV mPG (per 10 mmHg increase) | 0.95 (0.85–1.07) | 0.384 | 0.96 (0.84–1.10) | 0.568 |
| LVGLS (per 1% decrease) | 1.11 (1.06–1.17) | <0.001 | 1.08 (1.01–1.15) | 0.022 |
| AAS |  |  |  |  |
| Per 500-unit decrease | 1.46 (1.26–1.70) | <0.001 | 1.33 (1.12–1.58) | 0.001 |
| Per 1-SD decrease | 1.58 (1.32–1.88) | <0.001 | 1.41 (1.14–1.73) | 0.001 |
| Joint model of LVGLS and AAS† |  |  |  |  |
| LVGLS (per 1% decrease) | - | - | 0.97 (0.87–1.07) | 0.522 |
| AAS (per 500-unit decrease) | - | - | 1.44 (1.07–1.94) | 0.016 |

\* Each aortic valve disease or strain-based variable was evaluated in a separate multivariable Cox regression model adjusted for age, sex, LVEF, LVMI, RWT, LAVI, E/e', hypertension, diabetes mellitus, eGFR, hemoglobin, and albumin.
† The joint model included LVGLS and AAS simultaneously, with the same covariate adjustment, to assess their mutually adjusted associations with the primary outcome. AAS, afterload-adjusted strain; AR, aortic regurgitation; AV mPG, mean transaortic pressure gradient; AV Vmax, peak aortic jet velocity; eGFR, estimated glomerular filtration rate; E/e', ratio of early transmitral flow velocity to early diastolic mitral annular velocity; LAVI, left atrial volume index; LVEF, left ventricular ejection fraction; LVGLS, left ventricular global longitudinal strain; LVMI, left ventricular mass index; RWT, relative wall thickness; SD, standard deviation.

In separate analyses of the component outcomes, lower AAS remained independently associated with both all-cause death and HHF (**Tables S1** and **S2**). Among the 75 deaths occurring as the first primary composite event, 24 (32.0%) were cardiovascular and 51 (68.0%) were non-cardiovascular. Lower AAS remained independently associated with the cardiovascular-specific composite outcome of cardiovascular death or HHF (adjusted HR 1.61 per 1-SD decrease, 95% CI 1.20–2.15; p=0.001; **Table S3**). In a sensitivity analysis restricted to patients with at least mild AR (n=477), lower AAS remained independently associated with the primary outcome, both per 500-unit decrease in the raw value (adjusted HR 1.44, 95% CI 1.10–1.89; p=0.009) and per 1-SD decrease (adjusted HR 1.54, 95% CI 1.12–2.14; p=0.009) (**Table S4**). The association between lower AAS and the primary outcome remained consistent across sensitivity analyses accounting for AVR as a competing event, a time-dependent covariate, or a censoring event. A similar direction of association was observed in the post-AVR landmark analysis (**Figure S2**).

For the secondary outcome, a total of 120 events occurred. The cumulative event probability of AVR differed significantly across concomitant AR categories and according to LVH status, whereas no significant differences were observed according to RWT category or AAS status (**Figure 4B**). In Cox proportional hazards analyses, increasing AR severity, along with higher AV Vmax and AV mPG, was associated with a higher risk of the secondary outcome (**Table S5**). LVMI was also independently associated with AVR, whereas RWT and AAS were not. Consistent with these findings, restricted cubic spline analysis demonstrated an increasing risk of AVR with higher LVMI, whereas RWT and AAS showed no significant associations with AVR (**Figure S1B**).

### 3.3. Incremental Prognostic Value of AAS

The incremental prognostic value of AAS for the 1-year risk of the primary outcome was evaluated using three nested models. The addition of LVMI and RWT did not materially improve model performance. In contrast, the addition of AAS increased the 1-year AUC from 0.762 in Model 2 to 0.788 in Model 3 (p<0.001) and increased Harrell’s C-index from 0.727 to 0.744 (p<0.001). Continuous NRI also improved significantly (+0.425; p=0.020), whereas the increase in IDI was not statistically significant (+0.0120; p=0.200) (**Figure 6A**). Cox model–predicted 1-year risks increased progressively with decreasing AAS, with similar patterns observed in both sexes (**Figure 6B**). Decision-curve analysis further showed that Model 3 provided greater net benefit than Model 2 across threshold probabilities of 2% to 15% (**Figure S3**).

**Figure 6.**
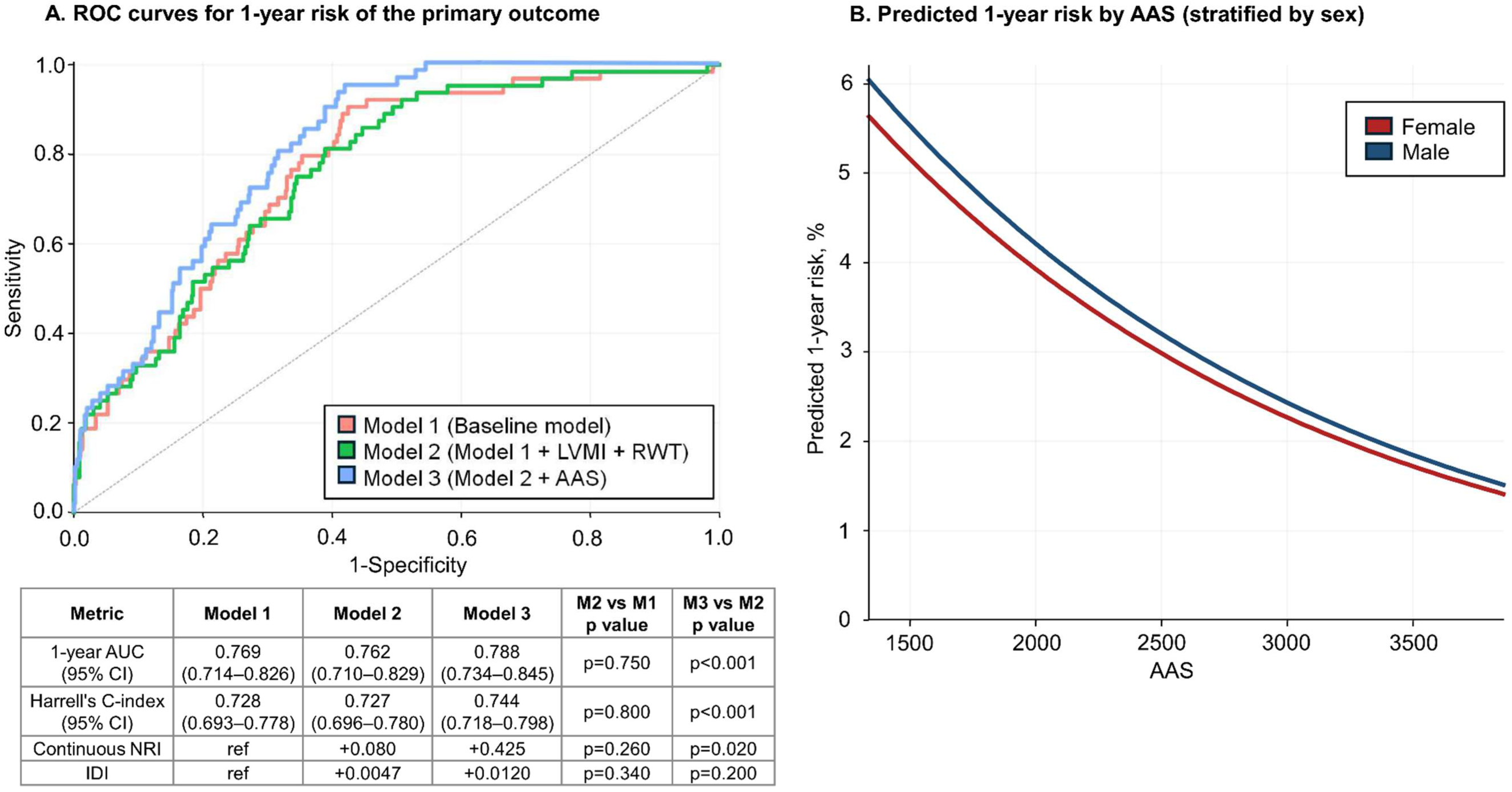
Incremental prognostic value of AAS. (A) Receiver operating characteristic curves for predicting the 1-year primary outcome. Model 1 included age, sex, LVEF, LAVI, E/e′, hypertension, diabetes mellitus, eGFR, hemoglobin, and albumin. Model 2 additionally included LVMI and RWT, and Model 3 additionally included AAS. The addition of AAS increased the 1-year AUC from 0.762 in Model 2 to 0.788 in Model 3 (p<0.001) and increased Harrell’s C-index from 0.727 to 0.744 (p<0.001). Continuous NRI and IDI were also evaluated. (B) Cox model–predicted 1-year risks of the primary outcome according to AAS in women and men. Continuous covariates were fixed at their cohort median values, and binary covariates were fixed at their most frequent categories. AAS = afterload-adjusted strain; AUC = area under the curve; eGFR = estimated glomerular filtration rate; IDI = integrated discrimination improvement; LAVI = left atrial volume index; LVEF = left ventricular ejection fraction; LVMI = left ventricular mass index; NRI = net reclassification improvement; RWT = relative wall thickness.

## 4. DISCUSSION

In this cohort of patients with moderate or greater AS across varying degrees of concomitant AR, we evaluated an integrated echocardiographic framework incorporating LV geometric remodeling and myocardial deformation in relation to the prevailing systolic pressure load. Three principal findings emerged. First, increasing AR severity was associated with progressive LV geometric remodeling, characterized by LV dilation, increased LVMI, and reduced RWT. Second, despite differences in LV geometry and transvalvular hemodynamics, AAS remained comparable across concomitant AR categories. Third, lower AAS was independently associated with the primary outcome and provided prognostic information beyond clinical and LV geometric variables.

### 4.1. Ventricular Remodeling Across Concomitant Aortic Regurgitation Severity

In isolated AS, chronic pressure overload promotes concentric hypertrophy, whereas isolated AR induces eccentric remodeling with LV dilation. In patients with AS, increasing degrees of concomitant AR introduce an additional volume-loading component that may modify the resultant LV geometric phenotype.^1–3^ The inclusion of patients without AR allowed the incremental ventricular changes associated with concomitant AR to be characterized across the cohort. In this study, increasing AR severity in the setting of moderate or greater AS was associated with a progressive shift toward a dilated LV phenotype, characterized by increased LVEDVi, increased LVMI, and reduced RWT. These findings demonstrate that conventional geometric measures capture complementary aspects of the ventricular response: LVMI reflects the magnitude of hypertrophic remodeling, whereas RWT characterizes the shift from concentric toward more eccentric geometry. Although LVMI was associated with a higher risk of AVR, neither LVMI nor RWT was independently associated with death or heart failure hospitalization. This divergence suggests that structural remodeling may more closely reflect the severity and progression of valvular loading, whereas adverse clinical outcomes may depend more directly on the accompanying impairment in myocardial performance.

### 4.2. Afterload-Adjusted Strain and Myocardial Performance

Although LVGLS is a sensitive marker of subclinical myocardial dysfunction in valvular heart disease, its load dependency may complicate interpretation under varying loading conditions.^12,14^ This limitation is particularly relevant in AS, where myocardial deformation is influenced by both systemic arterial pressure and the transvalvular pressure gradient.15 AAS was therefore constructed as an integrated pressure–deformation index combining LVGLS with estimated LV systolic pressure, approximated as the sum of SBP and AV mPG based on a previously described approach in patients with AS.^16^ Accordingly, AAS reflects myocardial deformation in relation to the prevailing systolic pressure load rather than strain normalized for afterload. In this study, despite increasing AR severity and higher transvalvular gradients, both LVGLS and AAS remained largely consistent across concomitant AR categories, suggesting similar myocardial performance in relation to systolic pressure load at the group level. However, the association between lower AAS and the primary outcome was directionally consistent across the no-AR, mild-AR, and moderate-or-greater-AR groups, with no evidence of interaction according to concomitant AR category. In addition, lower AAS remained independently associated with the primary outcome in the sensitivity analysis restricted to patients with at least mild AR, indicating that its prognostic association was preserved after excluding patients without AR. When AAS and LVGLS were included simultaneously in the multivariable model, AAS remained independently associated with the primary outcome, whereas LVGLS was no longer statistically significant. Although AAS mathematically incorporates LVGLS and this joint model should therefore be interpreted cautiously, these findings suggest that the prognostic information captured by AAS was not fully explained by LVGLS alone. Consistent with this interpretation, the association was directionally stronger in patients with reduced LVGLS, although the interaction according to LVGLS status was not statistically significant.

Several prior approaches have incorporated loading conditions into the assessment of myocardial performance in valvular heart disease. Valvuloarterial impedance reflects global LV afterload by integrating valvular and arterial loading but does not directly incorporate myocardial deformation.^15^ Pressure–strain loop–based indices provide a more comprehensive assessment of myocardial mechanics but require specialized analysis and should not be considered equivalent to the single-point AAS used in this study.^13^ In this context, AAS provides a pragmatic approach that integrates myocardial deformation with estimated LV systolic pressure using routinely available echocardiographic parameters.

### 4.3. Limitations of Valve-Centric Assessment

Conventional valve-centric parameters, including AV Vmax and AV mPG, are well-established measures of disease severity and prognostic markers in patients with AS.4 These indices quantify the pressure burden imposed by the stenotic valve and form the basis of current guideline-based assessment. However, in patients with concomitant AR, valve-specific parameters may incompletely characterize the ventricular consequences of combined pressure and volume loading.^5^ Moreover, valve severity alone may not fully capture the accompanying LV remodeling and myocardial dysfunction, which have important prognostic implications.^6–8^

In the present study, AV Vmax and AV mPG were associated with AVR but not with death or heart failure hospitalization. This divergence suggests that conventional valve parameters remain relevant to assessment of valvular severity and subsequent intervention, whereas the risk of adverse clinical outcomes may also depend on the ventricular response to chronic loading. Accordingly, complementary assessment of LV geometry and myocardial performance may provide additional information beyond valve severity alone.

### 4.4. Complementary Roles of LV Geometry and AAS in Risk Stratification

The findings of this study highlight the complementary roles of LV geometric remodeling and myocardial performance in patients with moderate or greater AS across varying degrees of concomitant AR. Conventional geometric measures, including LVMI and RWT, characterize structural adaptation to chronic loading conditions, whereas AAS reflects myocardial deformation in relation to the prevailing systolic pressure load. AAS was independently associated with the primary outcome and provided prognostic information beyond clinical and LV geometric variables. These findings underscore the distinction between structural remodeling and myocardial performance. Current clinical practice primarily relies on valve-centric parameters and LV remodeling to guide intervention, but does not directly account for myocardial dysfunction. Accordingly, incorporation of AAS may refine risk stratification by providing complementary information on myocardial performance beyond valve severity and LV geometry. This concept is consistent with emerging evidence supporting consideration of the ventricular response when evaluating the timing of intervention in AS.^16,17^ Taken together, an integrated assessment incorporating valve severity, LV geometry, and myocardial performance may provide a more comprehensive characterization of ventricular response than assessment based on a single domain.

### 4.5. Limitations

Several limitations should be acknowledged. First, this was a retrospective, single-center study, which may limit generalizability. External validation in independent populations is therefore required to establish the reproducibility and clinical applicability of AAS. Second, AR severity was assessed using an integrative qualitative approach and may be subject to interobserver variability. Third, approximately half of the patients had no concomitant AR, whereas only 11.9% had moderate or greater AR. Accordingly, comparisons across AR categories should be interpreted as characterizing the incremental effects of concomitant AR within an AS cohort rather than as definitive comparisons across established MAVD phenotypes. Further studies are needed to validate the prognostic value of AAS in populations with clinically significant MAVD. Fourth, AAS was derived using SBP and AV mPG as surrogates of afterload and does not fully capture arterial compliance or valvuloarterial impedance. Furthermore, because AAS is based on a single-point product of LVGLS and estimated LV systolic pressure, it should not be considered equivalent to pressure–strain loop–derived myocardial work.^13^ Direct comparison with formal myocardial work indices was not possible because dedicated software for pressure–strain loop reconstruction was unavailable in this retrospective dataset. Fifth, additional parameters such as natriuretic peptides and aortic valve calcium burden were not systematically available. Sixth, the timing of AVR as the secondary outcome may have been influenced by physician discretion and institutional practice. Although multiple sensitivity analyses were performed to account for AVR during follow-up, residual treatment-related modification of subsequent clinical risk cannot be excluded. Seventh, low-flow, low-gradient AS was not specifically characterized, although patients with LVEF <40% were excluded, limiting inclusion of the most common underlying phenotype. Finally, LVGLS was measured using two vendor platforms, and inter-vendor variability in absolute strain values cannot be fully excluded.

## 5. CONCLUSIONS

Among patients with moderate or greater AS across varying degrees of concomitant AR, ventricular geometry and myocardial deformation in relation to systolic pressure load represented complementary dimensions of LV assessment. AAS provided a measure of myocardial performance in relation to the prevailing systolic pressure load and was independently associated with death or HHF, with prognostic value beyond clinical and LV geometric measures. An integrated approach incorporating valve severity, LV remodeling, and myocardial performance may refine risk stratification and support more comprehensive clinical assessment.

## Data Availability

The data that support the findings of this study are not publicly available due to institutional review board restrictions intended to protect patient confidentiality. Deidentified data will be made available from the corresponding author upon reasonable request.

## Contributors

J.S.: Data Curation, Methodology, Formal Analysis, Writing – Original Draft.

J.P.: Conceptualization, Supervision, Clinical Interpretation, Validation, Investigation, Data Curation, Formal Analysis, Writing – Review & Editing.

M.B.: Investigation, Supervision, Annotation.

H.-M.C.: Investigation, Supervision, Annotation.

I.-C.H.: Investigation, Supervision, Annotation.

Y.E.Y.: Investigation, Supervision, Annotation.

G.-Y.C.: Investigation, Supervision, Annotation.

All authors read and approved the final version of the manuscript.

## Data Sharing Statement

The data underlying this study cannot be made publicly available because of ethical restrictions imposed by the institutional review board, as public disclosure could compromise patient confidentiality and privacy. Requests for access to a minimal anonymized dataset may be directed to the corresponding author.

